# Health System Surveillance to Advance Mental Health Parity for Children

**DOI:** 10.1101/2025.09.25.25336447

**Authors:** Darja Beinenson, Jacqueline Joyce, Hillary Henderson, Jeena Patel, Gracie Frericks, John N. Constantino

## Abstract

**Objective:** Parity between physical and mental health for insured individuals is protected by federal law but unfulfilled, in part, because deficiencies are notoriously difficult to document.

**Methods:** We implemented real-time, systematic clinical documentation of gaps in access to mental health services for an outpatient cohort representing 2,149 children discharged from Children’s Healthcare of Atlanta emergency departments for behavioral crisis May-Dec 2024.

**Results:** Sixty percent of patients discharged to the community with clear recommendations for expedited follow up were not receiving any service at 30–75-days. For those internally referred to the system’s outpatient psychiatric practice, the documentation system confirmed medical necessity and captured accessibility or non-accessibility for each element of evidence-based care of each patient’s treatment plan.

**Conclusion:** Feasible revisions of electronic health record documentation, coupled with clinical accountability pathways can enable health systems to assume responsibility for continuous, real-time epidemiologic estimation of gaps in the adequacy of provider networks to meet the mental health needs of insured children.

**Highlights:** *We modeled health-system-based surveillance of gaps in access to medically necessary mental health care for children, toward fulfillment of federal mental health parity law*.

*Health system-based monitoring of unmet needs was feasible and generated real-time epidemiologically-relevant estimates of gaps in community capacity for mental health services*

---

Federal mental health parity law (1) requires that if an insurance plan provides both medical/ surgical benefits and mental health/substance use disorder benefits, then the latter must not be more restrictive than the former. Gaps in parity have been notoriously difficult to document, which has compromised accountability and fulfillment of the law (2). One of the most salient drivers of non-fulfillment of mental health parity in the U.S. is so-called “network inadequacy” (3), which results from under-recruitment of mental health clinicians to participate in a provider network (i.e. to “accept the reimbursement” of a payor for their service), and results in either unmet need or decisions of adequately-resourced families to self-pay for service. In mental health, both occur at rates that are grossly disproportionate to that for physical health conditions. This is difficult to monitor for outpatient services in an insured population because outside of the monitoring of *denials* (which are common for high-cost, higher-intensity mental health services such as residential treatment, but uncommon for lower-cost, higher-frequency outpatient services) payors do not track what they have inadequate capacity to provide, and anecdotal patient and provider complaints cannot reliably estimate the magnitude of inadequacy for an insured population.

Principal manifestations of the consequences of broad failure to achieve mental health parity for children in the U.S include a) relative inaccessibility of mental health care (4), and b) the rate of self-pay, which is 20 times higher for outpatient mental health care than for outpatient medical care (5), summarized schematically in Figure 1. Both disproportionately affect lower-income families on governmental insurance plans (managed Medicaid programs) who are unable to afford out-of-network care. In a recent analysis of claims and State-level administrative data from Georgia, for example, 34% of the demand from Medicaid-insured children in Georgia remained unmet, and 25% of the Georgia census tracts (rural 79%; urban 16%) had less than 50% service coverage (6). Retrospective data such as these can take years to acquire and assemble, and because of turnover in client-plan relationships, service gaps identified by these methods may no longer be actionable by the time they are discovered.

**Figure 1.**
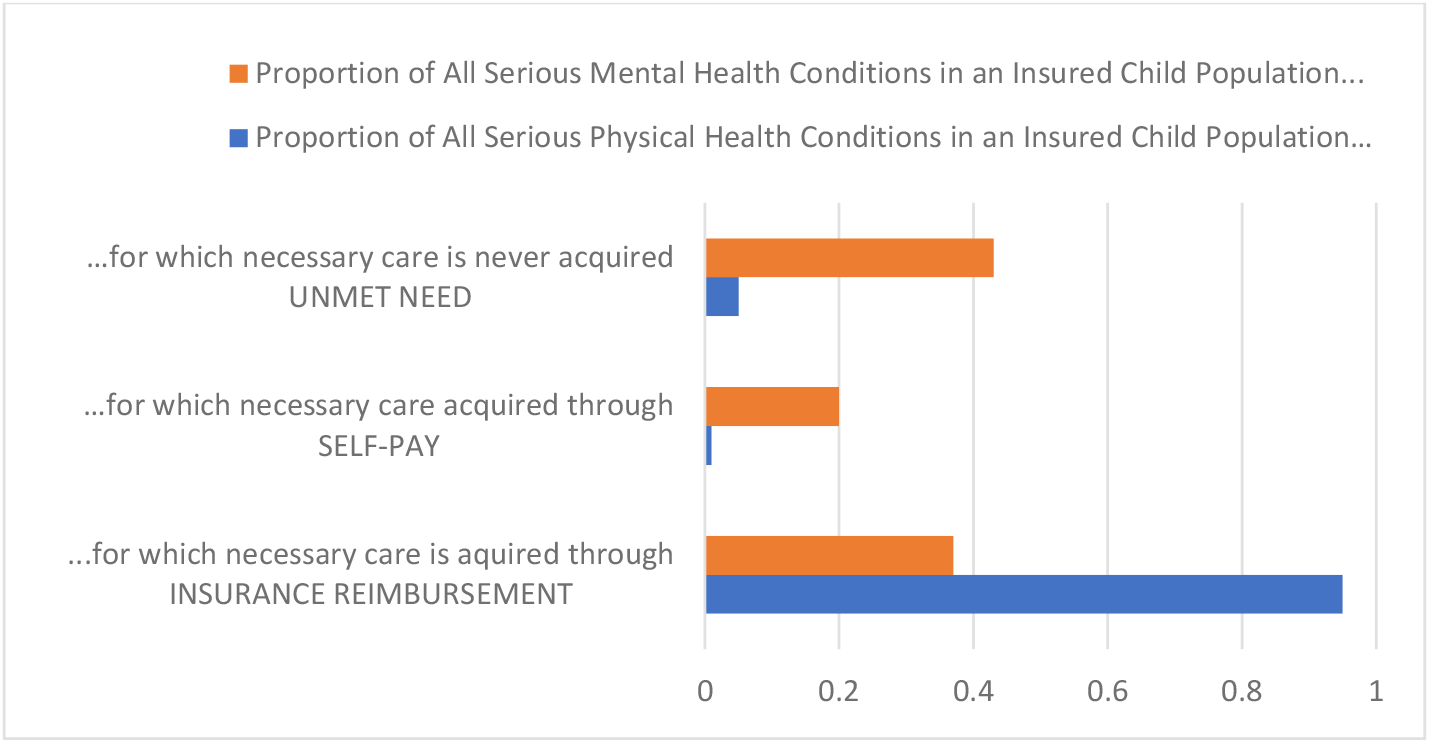
Depiction of three measurable indices of lack of parity between physical health and mental health care for children; inadequate clinical capacity within insurance provider networks is a major driver of these contrasts (see text for citations).

Here we summarize clinical quality data from systematic, real-time clinical documentation of mental health service gaps within a population-representative set of children in behavioral crisis in a large health system. We are not aware of any previous report of results of establishing such a system of documentation, and we communicate these results to validate a potentially scalable solution for detection and ultimately resolution of mental health parity gaps within insured populations. At the outset it is important to clarify that systematic documentation of such gaps requires three foundational elements of information (7), the combination of which is rarely systematically executed in pathways of mental health care: *i)* documentation which unambiguously specifies the medical necessity and timing of covered elements of care (the standard for defining and legally enforcing mental health parity)—an advantage for health systems is availability of physicians to assert medical necessity of an intervention; *ii*) subsequent ascertainment of whether the interventions specified in the treatment plan actually occurred--this is commonly an unknown when care is fragmented and/or there is no clinician accountable for an episode of mental health care; and if not (*iii)* whether the recommended service was or was not accessible to the patient within the provider network of the patient’s insurer (which requires direct inquiry of insurers to determine whether their provider network has adequate capacity to accommodate an appointment for the necessary service within a medically-acceptable timeframe). The latter typically requires care coordination by social workers or nurse case managers who are more commonly employed within health systems than in individual practices.

## METHODS

This report was developed through an analysis of data--STUDY00002509 determined non-research by Children’s Healthcare of Atlanta IRB--acquired in the course of clinical care for a population of youth presenting for primary behavioral concerns to the emergency departments of the three children’s hospitals in our system (Arthur M. Blank Hospital—formerly Egleston Children’s Hospital—Scottish-Rite Hospital, and Hughes Spalding Hospital), and/or a newly established outpatient Center (the Zalik Center for Behavioral and Mental Health) during the months from May through December of 2024. Because the three hospitals receive 25% of all emergency department encounters of adolescents in behavioral crisis in the State of Georgia, the E.D. cohort described above approximates an epidemiologic sampling frame.

In the first analysis, we reviewed outreach data on 132 children who were discharged from the emergency departments directly to the community (external to our own outpatient clinical program) over the surveillance period, selecting those whose E.D. records met the highest standard for informativeness of documentation of recommended mental health follow-up by emergency room physicians. Families were contacted 30-75 days following the E.D. encounter to determine the status-of-acquisition of recommended mental health services; 72% of the families were successfully reached and engaged in a follow-up telephone interview.

The second analysis afforded more detailed information on the unmet needs of a subpopulation *internally* referred (in contrast to community-referred) to a new outpatient program of the health system, which was designed to augment capacity for specific health-system-based mental health services for children in the region (see below). There, we simultaneously implemented detailed surveillance of the fulfillment of intervention plans for all enrollees, which included *i)* those referred from the emergency departments (therefore whose acquisition of care is most comparable to the emergency crisis patients referred to the community) and separately ii) those referred from medical specialists across the health system. Whenever specific treatment recommendations by child and adolescent psychiatrists or psychiatric mental health nurse practitioners were unfulfilled at the time of follow-up, a clinic social worker or nurse case manager was engaged to seek access to the necessary service and when unable to be accessed within the provider network for the patient, this was reported back to the medical provider and documented in a database by individual patient I.D., unfulfilled element of care, and payor. All data elements were subsequently incorporated into the electronic health record of the health system.

It is important to note that the outpatient clinic afforded subsidized capacity for all enrolled patients—irrespective of payor source--to receive two discrete elements of specialty mental health care, for which national and regional data had established that access for Medicaid-insured outpatients was either significantly constrained (child and adolescent psychiatry) or non-existent (dialectical behavioral therapy, relevant to recovery of suicidal patients). For E.D.-referred patients, since the number of available slots could accommodate only a fraction of all E.D. Behavioral Crisis patients who required psychiatric follow-up during the surveillance period, assignment into the outpatient program occurred essentially at random (referrals were only accepted when an appointment was available within two weeks of the E.D encounter). The subsidy afforded coverage of expenses for provider salaries, training, and startup costs that were in excess of what was recoverable from insurance revenue for their clinical service during the surveillance period. The payor mix for each respective subpopulation is reported in Table 1; a majority of the children of the entire cohort reported were governmentally insured.

**Table 1.** Demographic characteristics of the respective clinical cohorts. I. All patients discharged from Children’s Healthcare of Atlanta emergency departments to the community (i.e. external to Children’s) following presentation for a behavioral crisis from May to December 2024; A representative sub population whose records were fully informative and who were contacted in a quality improvement effort to determine if recommended interventions were being received; A contemporaneous cohort of children referred internally to the outpatient psychiatric service of Children’s Healthcare of Atlanta from the emergency department; IV. All patients in the new outpatient program

|  | Discharged from Emergency Dept (E.D.) |  | Follow-Up of Representative Sample of E.D. Patients Discharged to the Community |  | E.D. Patients Internally Referred to the Crisis Recovery Arm of a New Outpatient Program |  | All Patients in the New Outpatient Program |  |
| --- | --- | --- | --- | --- | --- | --- | --- | --- |
|  | N | % | N | % | N | % | N | % |
| **Total** | 2149 |  | 95 |  | 274 |  | 2161 |  |
| **Unmet Needs** | NA | NA | 57 | 60% | 9 | 3.3%* | 120 | 5.6%* |
| **Gender** |  |  |  |  |  |  |  |  |
| Male | 918 | 42.7% | 42 | 44.2% | 146 | 53.3% | 1006 | 46.5% |
| Female | 1231 | 57.3% | 53 | 55.8% | 128 | 46.7% | 1155 | 53.5% |
| **Age** |  |  |  |  |  |  |  |  |
| ≤12 | 997 | 46.4% | 25 | 26.3% | 131 | 47.8% | 1048 | 48.5% |
| 13-20 | 1152 | 53.6% | 70 | 73.7% | 143 | 52.2% | 1113 | 51.5% |
| **Race** |  |  |  |  |  |  |  |  |
| White | 700 | 32.6% | 34 | 35.8% | 91 | 33.2% | 1047 | 48.4% |
| Black / AA | 1249 | 58.1% | 58 | 61.1% | 167 | 61.0% | 914 | 42.3% |
| Other/Unknown | 200 | 9.3% | 3 | 3.1% | 16 | 5.8% | 200 | 9.3% |
| **Ethnicity** |  |  |  |  |  |  |  |  |
| Hispanic or Latino | 306 | 14.2% | 12 | 12.6% | 36 | 13.1% | 329 | 15.2% |
| **Payor** |  |  |  |  |  |  |  |  |
| Managed care / commercial | 267 | 12.4% | 26 | 27.4% | 93 | 34.0% | 864 | 40.0% |
| FFS Medicaid | 287 | 13.4% | 13 | 13.7% | 39 | 14.2% | 276 | 12.8% |
| Medicaid CMO | 1287 | 59.8% | 47 | 49.4% | 120 | 43.8% | 887 | 41.0% |
| Tricare | 36 | 1.7% | 1 | 1.1% | 5 | 1.8% | 46 | 2.1% |
| Self-pay | 272 | 12.7% | 8 | 8.4% | 17 | 6.2% | 88 | 4.1% |
| **Presenting Concern (E.D.)** |  |  |  |  |  |  |  |  |
| Suicidality | 975 | 45.4% | 16 | 16.9%** | 112 | 40.9% | NA | NA |
| Homicidality | 122 | 5.7% | 2 | 2.1% | 8 | 3.0% | NA | NA |
| Environmental crisis/exposure | 225 | 10.5% | 31 | 32.6% | 39 | 14.2% | NA | NA |
| Unexplained change in mental state | 350 | 16.3% | 23 | 24.2% | 72 | 26.3% | NA | NA |
| Concern for imminent risk of harm (non-suicidal) | 150 | 6.9% | 15 | 15.8% | 10 | 3.6% | NA | NA |
| Unknown | 327 | 15.2% | 8 | 8.4% | 33 | 12.0% | NA | NA |

*Note.* \*This percentage is following the provision of subsidized psychiatric care (medically-recommended and not previously established for the patients in these cohorts) and DBT-based crisis recovery intervention (medically indicated for 40% of the crisis recovery patients but inaccessible to governmentally-insured (Medicaid CMO) outpatients in our region. \*\*Suicidal patients were less common in the group discharged to the community; suicidal patients were more commonly either hospitalized or internally referred to the new outpatient program at Children's (which offered immediate access to psychiatric care and DBT-based therapy for recovery from suicidality).

## RESULTS

Among the 2149 children discharged from the Emergency Department (E.D.) during the surveillance period, 12% had recurrent visits for behavioral crisis, and among these 20% were documented by mental health specialists never to have received outpatient mental health services at either the index visit or subsequent visits. For a representative sample of the entire group, outreach workers confirmed that most (60%) were not receiving *any* specific recommended mental health service at 30-75 days follow-up. Demographic characteristics and proportions of patients with unmet needs for each group are summarized in Table 1.

Psychiatric evaluations of internally referred E.D. behavioral crisis patients confirmed that a comparable majority required either a) child psychiatric care that had not been previously established in the community and/or b) dialectical behavioral therapy-based intervention for active suicidal symptomatology (40.9% of the ED referred group), for which we were unable to identify any provider accepting governmental-insurance for outpatients in the community at the outset of the surveillance period. Residual unmet needs of the patients included cognitive behavioral therapy, intensive outpatient programming, applied behavior analysis-based intervention, and other specialized psychosocial treatments. In our population, social work care coordination incurred an average of two hours of effort per patient; it resulted in successful identification of treatment resources for many patients, recognition of family non-compliance for some, and for others confirmed inadequacy of the insurer’s provider network to materialize a timely appointment for a necessary service (here reported as unmet needs), all of which represent valuable outcomes of care coordination.

## DISCUSSION

These health system data converge on a rate of current community capacity for the fulfillment of intervention plans of youth in crisis that is less than half of what would be medically-indicated for their conditions, and are in keeping with data from recent retrospective studies of Medicaid-insured patients in Georgia (6). Medical providers demonstrated the feasibility of documenting the fulfillment or non-fulfillment of their own treatment recommendations during clinical follow-up encounters. Gaps in insurance provider network capacity observed here— substantially resolved within our health system based on a subsidy to recruit clinicians into a dedicated outpatient service—have been similarly documented in explorations of provider capacity within regional populations of insured patients using “secret shopper” methodology (9).

Implementing systematic documentation among outpatient clinicians was straightforward, involving (a) discrete-field entry of recommendations for medically necessary intervention in each evaluation, (b) unequivocal ascertainment of whether the necessary elements of care were acquired in each follow up encounter, and for unmet needs (c) to determine whether residual unmet needs resulted from inadequacy of the insurer’s provider network to materialize a timely appointment for the necessary service or from other reasons.

## CONCLUSIONS

Health systems have unique capability to systematically document access to mental health care for large numbers of patients who are epidemiologically-representative of the populations of their respective payors. A distinct advantage over anecdotal reports or retrospective analyses of claims data is that the information in this approach can be acquired naturally in the course of clinical care, in real time, and is directly auditable by patient, treatment type, and payor through the electronic health record. Given the ongoing toll of a youth mental health crisis (10), that these data pertaining to youth in crisis reflect serious gaps in access to necessary intervention services, that mental health parity is a matter of federal law, and that the intention to enforce mental health parity through implementation of complaint portals has largely failed, we conclude that health systems should consider assuming responsibility for systematic documentation of mental health service gaps for their patients. Such documentation carries the promise of real-time accrual of information on actionable service needs that can be targeted and resolved through coordinated efforts of health systems and insurers.

## Data Availability

Clinical data incorporated into this analysis are not publicly available at this time.

## Abbreviations

E.D.: Emergency Department
PMHNP: Psychiatric Mental Health Nurse Practitioner
MD: Doctor of Medicine.

## Disclosures and Acknowledgments

The authors warrant that there are no financial relationships with commercial interests of relevance to this report. This research was supported by a gift from the William Randolph Hearst Foundation to Children’s Healthcare of Atlanta

## Acknowledgments

Funding for the clinical team engaged in supplemental outreach services described in this report was provided by a gift from the William Randolph Hearst Foundation. The authors gratefully acknowledge the work of the clinicians who generated the clinical data for documentation of unmet mental health needs of their patients. The authors also acknowledge Tammy Bamlett Sherman, MBA, MHA, CMPE, Megan Costa, Lisa Schneider, MS, BSN, RN, PMH-BC, NEA-BC, Kaitlin Smith, MHA, PMP, and the administrative team of the Center for Behavioral and Mental Health of Children’s Healthcare of Atlanta for their assistance in organizing the clinical program data that enabled this analysis

## Data Sharing Statement

This analysis was conducted using anonymized clinical program data, with access authorized for quality improvement purposes. The original patient health information data is not available to be shared.

